# Genetic Counseling Educational Videos Significantly Improve Access to Genetic Testing and Counseling for Inpatients with Cardiovascular Disease

**DOI:** 10.64898/2026.06.24.26356505

**Authors:** Emily E. Brown, Bryana Rivers, Jeff Day, Lisa R. Yanek, Kathryn Nunez, Catherine Gordon, Crystal Tichnell, Rebecca McClellan, Andreas Barth, Amy Sturm, Carolyn Applegate, Cynthia A. James, Brittney Murray

## Abstract

**Background:** Genetic testing for inherited cardiovascular conditions is recommended by multiple national guidelines to inform medical management. However, access to genetic counseling and testing is often limited particularly in the inpatient setting. Cardiologists cite lack of access to genetic counselors as a reason for not pursuing genetic testing. Genetic test education videos have been successfully implemented in the outpatient setting to increase patient volumes and decrease wait times, but they have not been studied in the inpatient setting.

**Methods:** This was a pilot study utilizing a genetic test education video in lieu of a traditional genetics consult for inpatients with a suspected hereditary cardiomyopathy, arrhythmia or familial hypercholesterolemia. Patients completed the cardiovascular multidimensional model of informed choice (MMIC) and SURE scales to assess whether the video provided sufficient information to make an informed decision.

**Results:** Over 22 months, 186 adult cardiovascular inpatients at Johns Hopkins Hospital were referred with 112 patients consenting to the study. Implementation of the pre-test videos increased the average number of monthly consults from 1 to 8. After watching the video, 90% (100/111) of patients made an informed choice to proceed with genetic testing using the MMIC scale. Using the SURE scale, 87% (97/112) of patients had no decisional conflict regarding whether or not to proceed with genetic testing.

**Conclusions:** These results indicate a pre-test genetic educational video could be considered as a strategy to scale access to genetic counseling and cardiovascular genetic testing, especially in hospital systems where access to genetics is limited in the inpatient setting.

## Introduction

Genetic testing for inherited cardiovascular conditions is recommended by multiple national guidelines to inform medical management.^1-3^ However, access to genetic counseling and testing is often limited potentially leading to missed diagnoses and mismanagement.^4,5^ In a retrospective review of a medical health record dataset containing 1.7 million patients, only 1% of individuals with a cardiomyopathy underwent genetic testing, far below the expected frequency of inherited cardiomyopathies.^2,6^ While there are many possible reasons for this discrepancy in genetic testing uptake, access to genetic counselors specifically is often cited by cardiologists as one of the rate limiting factors to incorporation of genetics into their clinics.^7,8^ Additionally, genetic counseling services are more routinely outpatient, whereas inpatient genetic counseling is reported by only 2-3% of genetic counselors.^9^ Incorporating genetic counseling and testing while inpatient may improve uptake primarily due to the ease of referral, convenience for patients, and less financial barriers for testing coverage.^10,11^However, inpatient genetic counseling consults are often very time intensive for providers and as a result the frequency of these consult is often limited.^12^

One possible solution bridging this gap of genetic counselor availability for inpatient consultation is implementation of genetic test educational videos. Such videos have been studied within the outpatient oncology genetics field for over two decades and have been found to increase the number of patients who received both genetic counseling and testing within a timely manner.^13-17^ Additionally, genetic test educational videos were shown to be non-inferior with respect to patients’ satisfaction between those who received in-person pre-test counseling versus those who watched a video.^14^ There is also evidence suggesting videos improve comprehension compared to baseline knowledge.^17,18^ Pre-test genetic counseling videos have not been studied in the inpatient setting or routinely in the cardiovascular setting, but previous research suggests that they could have similar outcomes.^19^

Therefore, to explore the utility of this strategy in the cardiovascular setting, an implementation study was designed utilizing genetic test educational videos for adults admitted with a suspected inherited cardiomyopathy, arrhythmia, or familial hypercholesterolemia.

## Methods

This study was approved by the Johns Hopkins School of Medicine institutional review board (IRB00420074) and participants gave written informed consent. The data that support the findings of this study are available upon request.

### Study Design

Eligible patients were referred by their inpatient team at Johns Hopkins Hospital from March 2024-December 2025, and consent was subsequently obtained by the study team during the patients’ admission after review of the patient’s medical history. Consented patients watched the genetic counseling pre-test video on an iPad provided by the study team. Patients then decided whether to proceed with genetic testing while admitted. For those who proceeded with genetic testing, the patient’s chart was reviewed by a genetic counselor and an appropriate genetic test was ordered. A post-test genetic counseling appointment was scheduled to review results and obtain a complete family history. After testing decision, participants were administered two surveys on the iPad to assess decisional conflict and informed decision making. Outcomes measures were based on the RE-AIM implementation science framework. ^20^

### Eligibility Criteria

Adult patients admitted to Johns Hopkins Hospital with a known or suspected inherited cardiomyopathy, arrhythmia, or familial hypercholesterolemia were eligible for the study. Exclusion criteria included (1) patients younger than 18 years old, (2) patients who previously underwent cardiovascular genetic counseling, (3) non-English speaking patients, (4) patients who were pregnant, (5) patients who were incarcerated (6) patients who were unable to provide consent. Eligible patients were identified by their inpatient team and referred to the study team who provided subsequent eligibility review.

### Participant Demographics

Participant demographics were obtained from the patient’s electronic medical record including race, biological sex, age, and diagnosis.

### Video Development

Previously developed pre-test genetic counseling videos for cardiomyopathies, arrhythmias, and familial hypercholesterolemia were modified for the inpatient setting.^21^ Video content was created by genetic counselors specialized in cardiology, based on information covered in a traditional genetic counseling session. Topics covered included basic genetic concepts, possible results and implications, cascade screening and familial risk, and genetic discrimination. One video was specifically developed for familial hypercholesterolemia and the other video discussed testing for both arrhythmias and cardiomyopathies. The videos were previously tested in populations with and without familiarity of inherited cardiac disease and increased knowledge and received high engagement scores.^21^

### Outcomes Measurements

Systems outcomes were assessed by comparing to historical data and current wait times. Historical inpatient volumes were calculated using the inpatient genetic counseling consult volumes from six months prior to the beginning of the study. Current wait times were calculated based on the date of referral and date of the appointment.

Two surveys were administered to participants: the multidimensional model of informed choice (MMIC) and SURE scale. The MMIC is a validated measure to assess if patients are provided with sufficient knowledge to make informed decisions about genetic testing. The measure consists of eight true/false knowledge questions and five Likert-scale questions regarding attitudes toward genetic testing.^22^ We used version 1 of the recently validated “cardiac genetic testing” measure for participants with cardiomyopathy or arrhythmia.^23^ For familial hypercholesterolemia, we used the FH MMIC knowledge scale which has a single battery of true/false questions.

Validation of the FH MMIC scale was recently completed with publication pending (personal communication, S. Christian). Individuals who answered at least 6/8 questions correctly were considered knowledgeable, and an attitude score ≥11 indicated a positive attitude (personal communication, S. Christian). Both MMIC measures are available in the Supplemental Materials. The SURE scale is a four-question validated scale to measure decisional conflict regarding proceeding with genetic testing (Supplemental Material). A score of 4/4 indicated no decisional conflict.^24^

### Statistical Analysis

Descriptive statistics were used to describe the study cohort’s demographics and indication. Quantitative statistics including Chi-squared or Fisher’s exact tests as appropriate were used to test associations of clinical and demographic variables with extent of knowledge, informed choice, and decisional conflict. Historic average consults per month were compared to consults during the study period using Mood’s median test. Analyses were conducted in SAS (v9.4, SAS Institute Inc., Cary, NC) and SPSS (version 28; SPSS Inc., Chicago, IL). A p value <0.05 was considered statistically significant in all analyses.

## Results

Over 22 months, 186 adult inpatients at Johns Hopkins Hospital were referred with 112/158 (71%) eligible patients consenting to the study (Figure 1). The majority of participants were male (68%, 77/112) and between the ages of 30-59 (60%,67/112). Over half of the individuals were Black (52%, 58/112). The majority of patients were diagnosed with a cardiomyopathy and/or heart failure (79%, 88/112) with a smaller proportion having a known or suspected inherited arrhythmia (18%, 20/112). Only three individuals were referred for familial hypercholesterolemia (Table 1). After watching the video two patients declined to proceed with genetic testing.

**Table 1.**
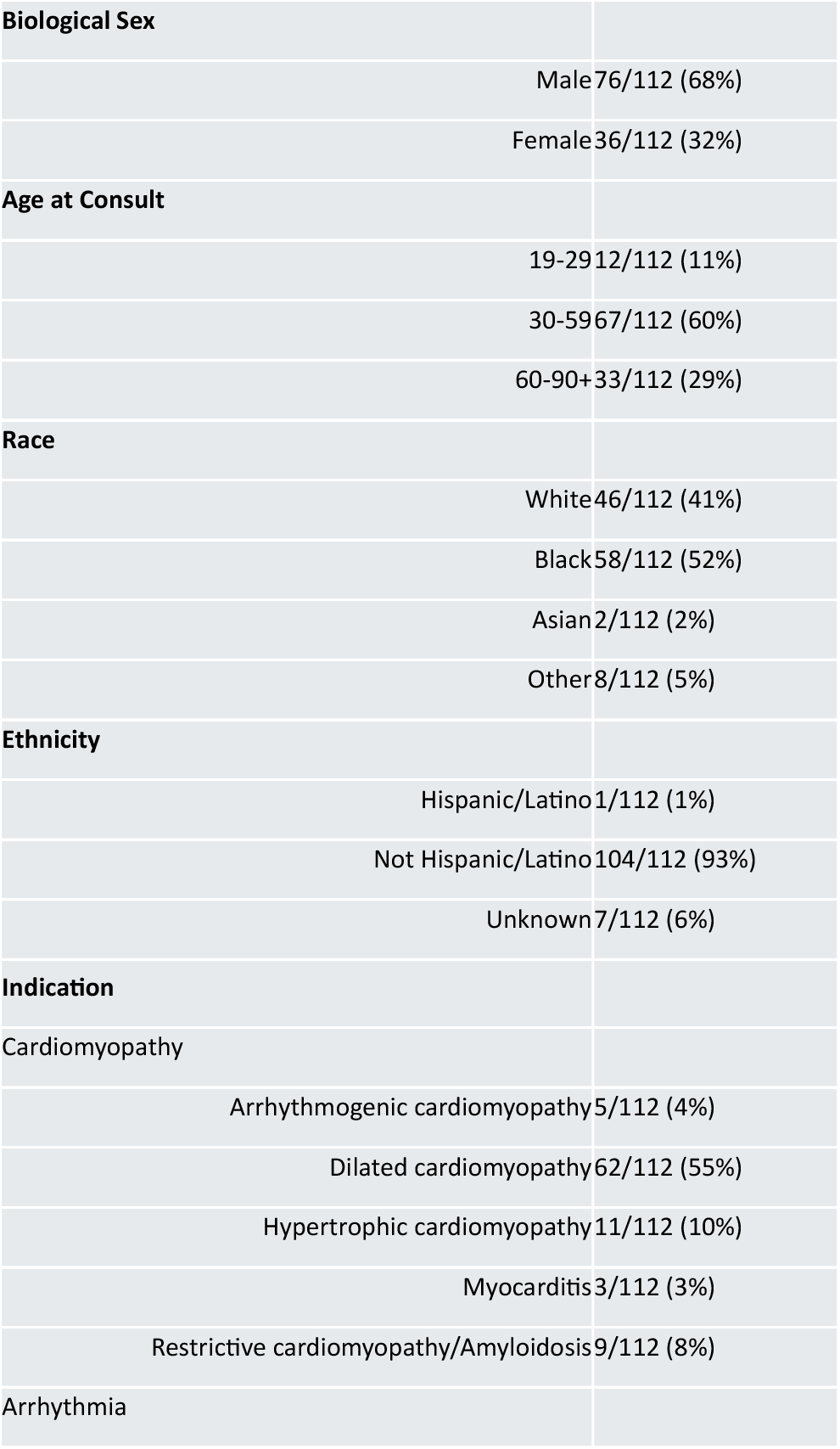
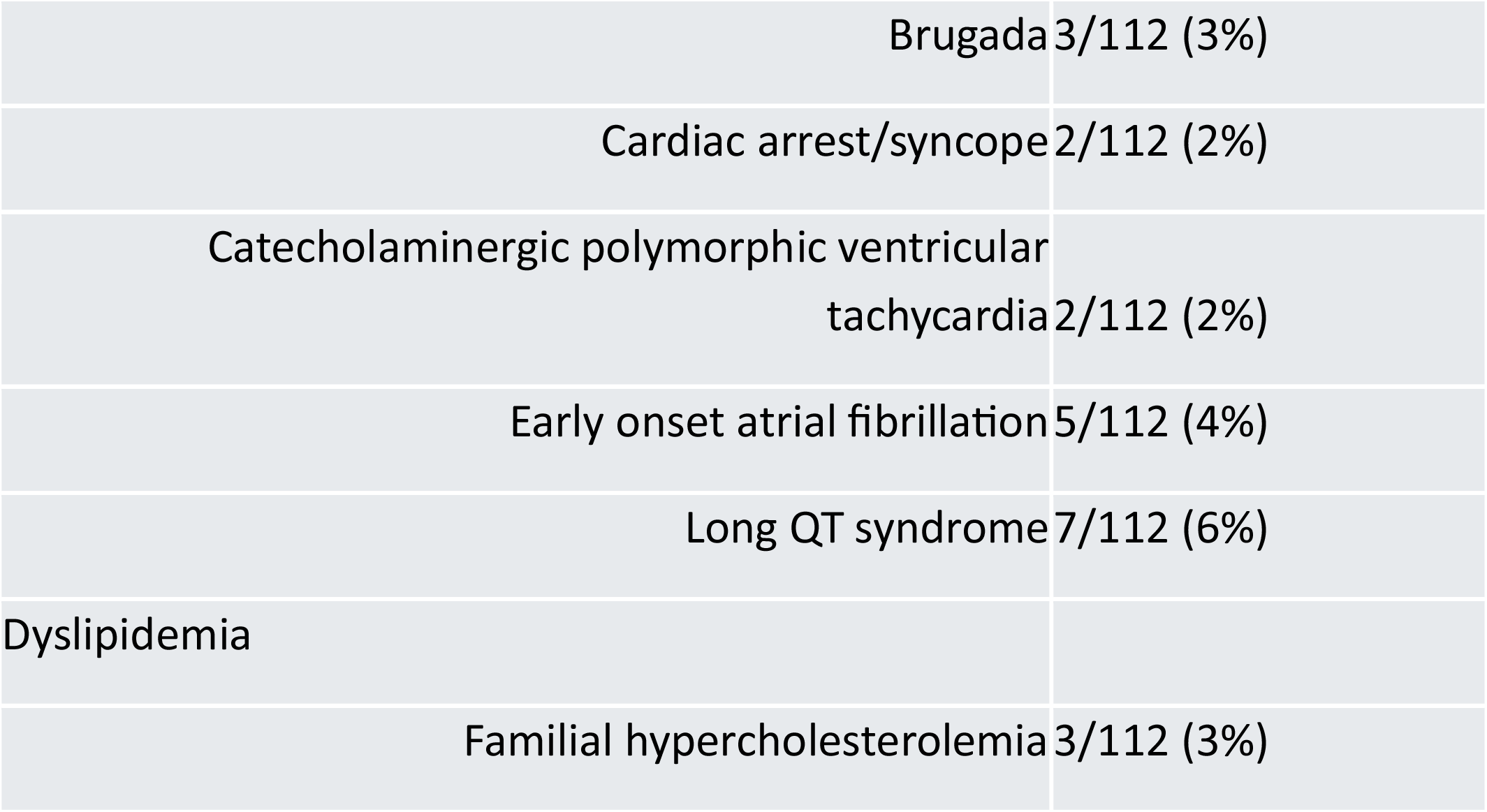
Patient demographics. Other= Pacific Islander, American Indian, or not listed.

**Figure 1.**
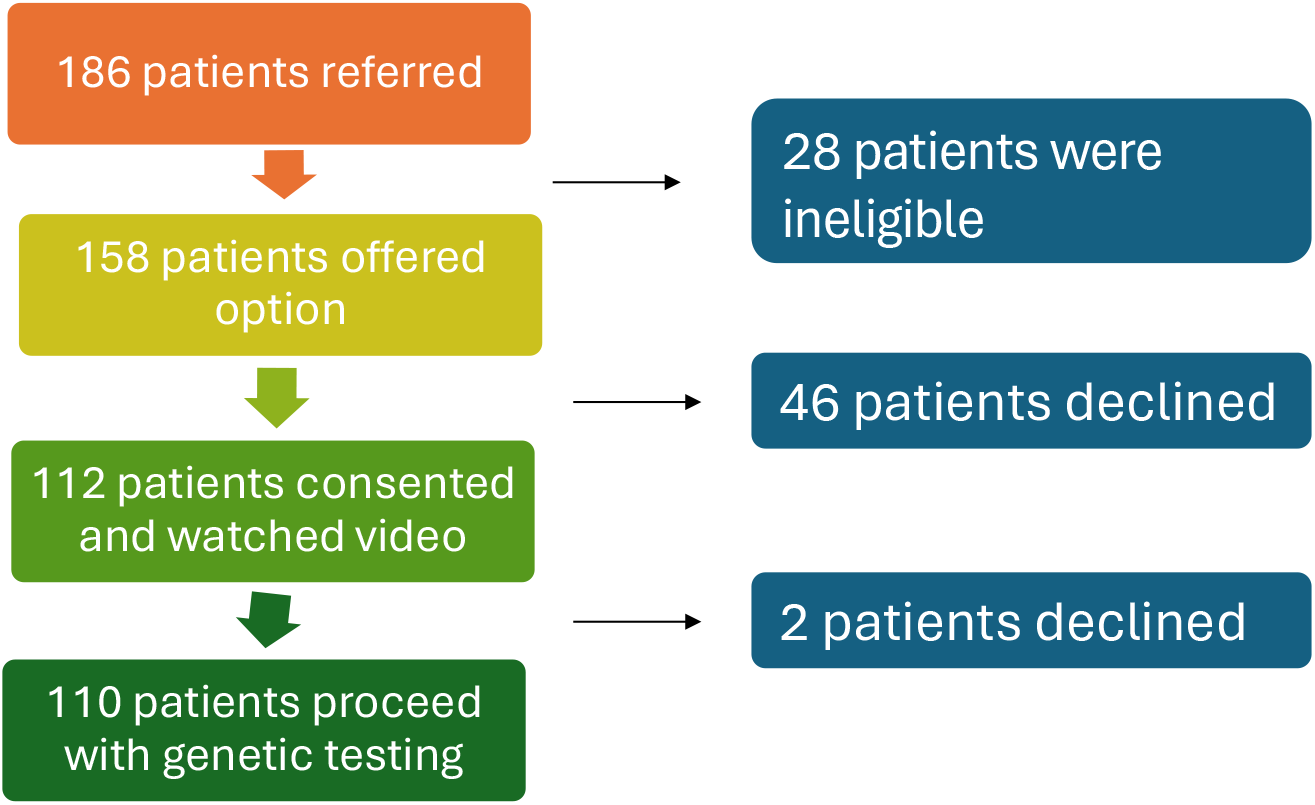
Diagram depicting enrollment.

### Increased Access

Historically, one (range 0-2) adult inpatient consult was completed per month on average. With the implementation of the genetic counseling pre-test video, the average number of consults increased to eight per month (range 2-15). In general, the number of consults per month increased over the study period with the average number increasing to ten per month over the last year of the study which is significantly higher than historical volumes (p=0.0043).

Consented patients saw a decreased wait time to results compared to outpatients. The average wait time during the study period to see a cardiovascular genetic counselor for an outpatient appointment at Johns Hopkins is 79 days (±52). Genetic test results typically take an additional 2-4 weeks to receive after the visit. Hence, it takes approximately 10-12 weeks from the time of referral for a patient to receive genetic test results outpatient. Having access to the pre-test genetic counseling video while admitted reduced the time to genetic test results to an average of 20 days (±6 days).

### Informed Choice

MMIC survey completion was 99% (111/112). Based on the MMIC, 93% (103/111) individuals were considered knowledgeable about genetic testing for cardiovascular disease after watching the video. Of the individuals who were identified as knowledgeable, 99 (96%) had a positive attitude about genetic testing and proceeded with testing. Greater than 95% of participants (107/111) stated they felt sure they made the best choice for them, and almost 95% (106/111) stated they had enough support and advice to make a choice after watching the educational video. Notably, there were no statistical differences between individuals categorized as knowledgeable after watching the video based on biological sex (p=0.44), age (p=0.69), or indication (p=0.84). There were differences based on race. All of the individuals who identified as White (n=45) were considered knowledgeable based on the questionnaire (p=0.02), and six out of 58 individuals who identified as Black did not answer enough questions correctly to be considered knowledgeable after watching the video (Table 2).

**Table 2.**
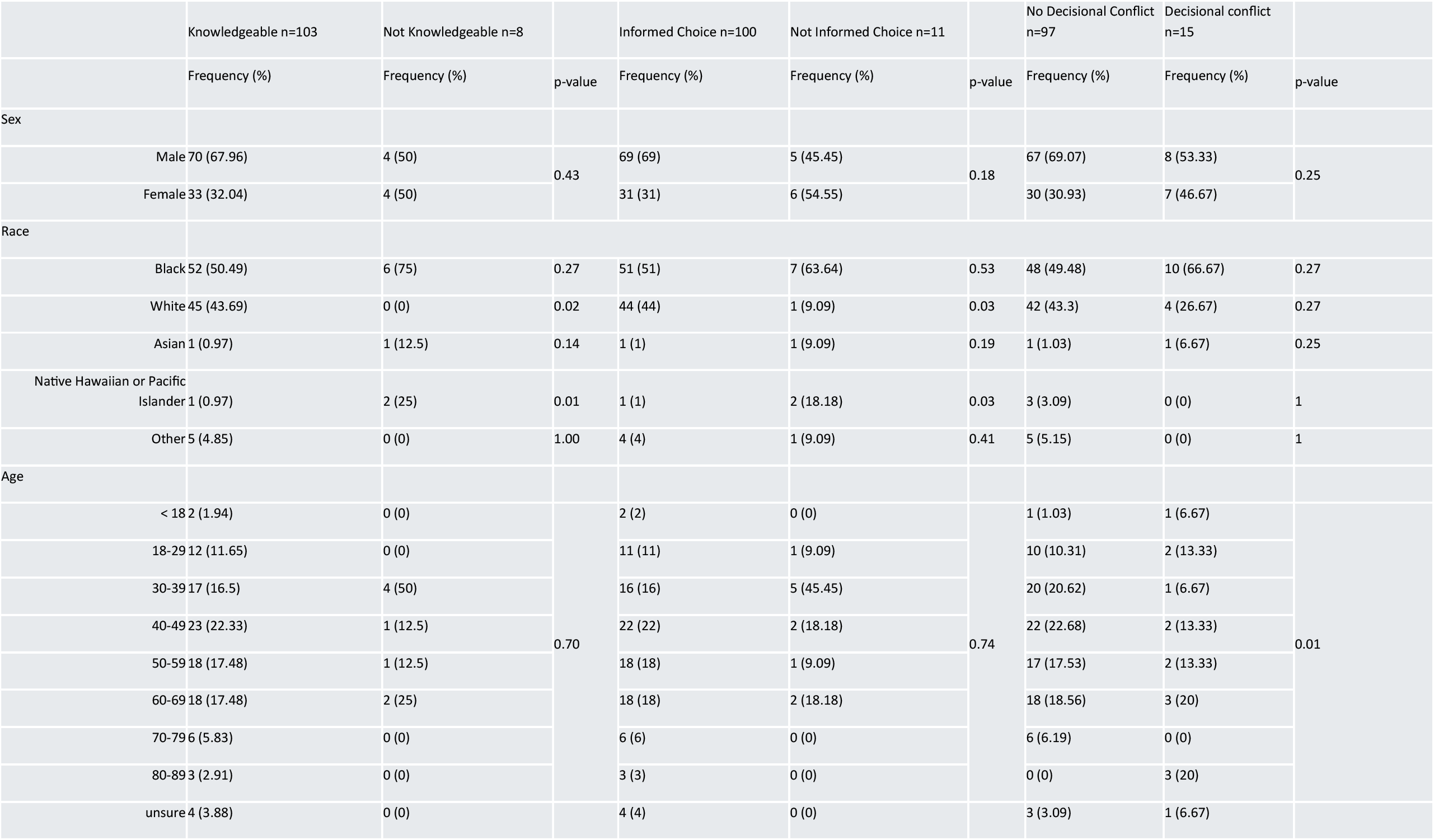
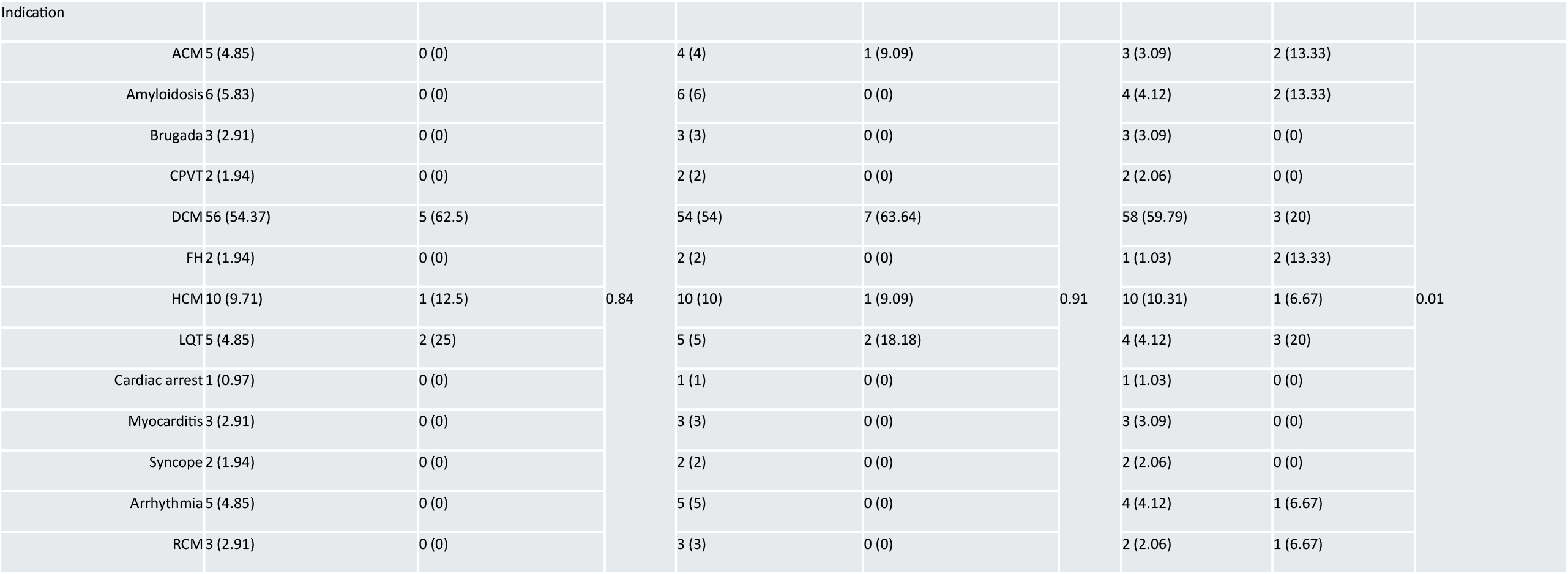
Statistical comparison of knowledge, informed choice, and decisional conflict based on biological sex, race, age, and indication. ACM= arrhythmogenic cardiomyopathy, CPVT= catecholaminergic polymorphic ventricular tachycardia, DCM= dilated cardiomyopathy, FH= familial hypercholesterolemia, HCM= hypertrophic cardiomyopathy, LQT= long QT syndrome, RCM= restrictive cardiomyopathy.

Three individuals had a negative attitude towards genetic testing but still proceeded with the test, and one individual had a negative attitude and declined testing. Overall, 90% (100/111) made an informed choice. There were no significant differences between individuals who made an informed choice and those did not based on biological sex (p=0.17), age (p=0.75), and indication (p=0.91). There were statistical differences based on race. Seven of 58 individuals who identified as Black did not make an informed choice (p=0.027), and only one individual who identified as White made an uninformed choice (p=0.02) (Table 2).

### Decisional Regret

All participants completed the SURE survey (112/112). Overall, 87% (97/112) individuals indicated they had no decisional conflict regarding whether to proceed with genetic testing after watching the video. Notably, 95% (106/112) of individuals indicated that they had enough support and advice to make a choice, and 96% (107/112) indicated they felt sure they had made the best choice for them after watching the video. There were no statistical differences based on race or biological sex (p=0.25) between those who had decisional conflict (Table 2). There were significant differences based on age (p= 0.01) and indication (p=0.0075) (Table 2).

### Follow-Up

Results were disclosed while the patient was still admitted 15% of the time (16/110). Five patients died before results were available. For the remaining individuals, a letter was sent or follow-up visit was scheduled to review the results. An outpatient visit was scheduled for 40 individuals to review their results (36%). A quarter of these individuals (10/40) no-showed for their follow-up visit.

## Discussion

The importance of genetic counseling and pre-test education accompanying genetic testing is highlighted in multiple national guidelines.^1-3^ However, despite these recommendations, there is often limited utilization of genetic testing and limited use of genetic counseling services.^9^ The are multiple reasons for this including limited provider availability for inpatient consults and challenges with billing and reimbursement for inpatient services.^9,12^ Given these challenges, we developed an educational video substitute for a traditional pre-test genetic counseling visit to try to increase access to these guideline-driven genetic test results for inpatients.

Overall, this study showed implementation of a pre-test genetic educational video for adult inpatients suspected of having an inherited cardiovascular condition successfully increased consult volumes, successfully increasing access to both pre-test genetics education and genetic testing. Utilizing the video increased the consult volume on average by 800%. Rather than being limited by genetic counselor availability, the number of consults seen reflected referral volume. While video creation has upfront costs, compared to hiring additional staff, this service delivery model may be more cost effective. Therefore, implementation of pre-test educational videos could be a viable alternative for some hospitals and clinics seeking to increase access to genetic counseling and testing in a guideline compliant but cost effective manner.^25^

Previous studies in oncology genetics have found pre-test videos to be effective methods of communication,^14,26^ and our study found similar results. In this study 90% of patients indicated they made a knowledgeable choice and 87% indicated no decisional conflict. Therefore, the data would suggest that the videos provided sufficient information for the majority of patients to make an informed decision which aligned with their values regarding genetic testing. Notably, there were a few patients who proceeded with genetic testing either without sufficient knowledge or when it did not align with their values. While this was a small number of patients, it suggests that pre-test counseling is not a “one size fits all approach” and some patients may be better suited for a discussion with a genetic counselor. Studies may further explore how to best identify those that may be best served with a pre-test genetic counseling visit; however, we are unable to explore in this study given the small dataset. An alternative approach may be for genetic counselors to work closely with inpatient teams to identify patients who may benefit from additional discussion.

Given the paucity of genetic counselors who practice in the inpatient setting^9^, these data suggest pre-test education videos can be an effective way to increase access to genetic counseling and testing for inpatients. Some patients may require follow-up with a genetic counselor prior to test ordering, especially if the inpatient team feels they have limited understanding.

### Limitations

It is important to acknowledge the limitations of this study. First, results come from a single tertiary hospital, and patient referral depended on discretion of the inpatient provider team which may have biased the participant characteristics, especially testing indication. In the systems outcomes analysis, the historical data is limited to only six months which limits comparisons. Additionally, when comparing outpatient appointment availability, this could vary based on many factors including urgency, patient preference/availability, and referral indication impacting the time to results.

## Conclusions

These results indicate a pre-test education video could be considered as a strategy to scale access to genetic counseling and testing especially in hospital systems where there are barriers to genetics services in the in-patient setting. For the majority of patients, the pre-test educational video provided sufficient information to allow them to make an informed decision regarding whether or not they would like to proceed with genetic testing.

## Data Availability

Data will be available upon reasonable request.

